# The impact of armed conflict on the prevalence and transmission dynamics of HIV Infection in Libya

**DOI:** 10.1101/2021.09.20.21263809

**Authors:** Mohamed Ali Daw, Abdallah Hussean El-Bouzedi, Mohamed Omar Ahmed

**Author notes:** Corresponding author: Mohamed A Daw, *MD, FTCDI*, Professor of Clinical & Microbial Epidemiology and Acting Physician of Internal Medicine, Department of Medical Microbiology & Immunology, Faculty of Medicine, University of Tripoli, Libya.

## Abstract

The interrelationships between HIV/AIDS and armed conflict are a complex phenomenon and studies are rarely devoted to this area of research. Libya is the second-largest country in Africa that has been evoked with war since NATO intervention in 2011. The country has also experienced one of the largest HIV outbreaks associated with the Bulgarian Nurses saga. The effect of the armed conflict on the dynamic spread of HIV is not well known. The objectives of this study were to determine the impact of armed conflict on the epidemiological situation of HIV infection in Libya and analyze the transmission dynamics of HIV strains during the conflict. We investigated the movement of HIV-infected people during the Libyan armed conflict and analyzed the HIV subtypes reported from 2011 to 2020 and followed up the infected cases all over the country. The patterns of HIV spread within the Libyan regions were traced and risk factors were determined during the conflict period. A total of 4539 HIV/AIDS patients were studied from the four regions during the Libyan conflict. Our data analysis indicated that Benghazi the biggest city in the Eastern region was the significant exporter of the virus to the rest of the country. Viral dissemination changes were observed within the country particularly after 2015. A major virus-flow from the Eastern region during the armed conflict associated with internally displaced people. This resulted in a dissemination of new HIV strains and accumulations of HIV cases in Western and Meddle regions. Although, there were no significant changes in the national prevalence of HIV/AIDS. Our data highlights the factors that complicated the spread and dissemination of HIV during the armed conflict which provides a better understanding of the interaction between them. This could be used to plan for effective preventive measures in tackling the spread of HIV in conflict and post-conflict settings.

## BACKGROUND

Human Immune Deficiency Virus (HIV) remains to be a serious problem, particularly within developing countries. Many of the countries affected by HIV are also ravaged by decades of wars and political instability. The interrelationships between armed conflicts and Infectious diseases have been considered inconsistent and studies rarely devoted to these phenomena (1). Armed conflicts and diseases usually link to one another in a positive direction, with violent conflict driving the incidence and prevalence of disease(s) upward. Infectious diseases vary greatly in transmissibility during armed conflicts. Airborne and fecal-oral pathogens such as tuberculosis, cholera, influenza, SARS-CoV 2(COVID-19 pandemic), smallpox, typhus, dengue fever, malaria, plague, and yellow fever can spread easily in conflict situations due to poor sanitation and population movements or refugee flows (2,3). This, however, is not the case of HIV which requires quite specific conditions for transmission. Therefore, studies are needed to highlight the impact of armed conflict on the epidemiological situation of HIV in war-torn countries (4).

Northern African region continues to account for the large majority of the world’s HIV/AIDS reported cases in the last decade and it was one of the regions that have been more acutely affected by large-scale violent conflict than other regions in the world (5). Interestingly no studies have been published about the dynamics of the relationship between the two crises. However, epidemiological studies in this area of research are limited and even, indicating an ambiguous and complex relationship between conflict and HIV prevalence levels worldwide. Studies from sub-Saharan Africa showed that at the end of the Angola conflict in 2002, the country’s HIV prevalence was relatively lower than the rates in other Southern African countries, which suggests that conflict may have slowed HIV spread in this case. Hence then, further studies are needed to highlight the dual-burden of violent conflict and infectious disease (6,7).

Libya, the second-largest country in Africa with the longest coast in the Mediterranean basin has been evoked in a major destructive armed conflict since 2011. During the conflict, health care services were continuously interrupted and the conflict caused massive internal population displacement. Over 1.5 million people have been internally displaced in Libya out of a total population of 6 million (8-10). Furthermore, the country has experienced one of the largest HIV/AIDS outbreaks associated with the Bulgarian Nurses saga in Benghazi in the Eastern region of the country (11,12). Suggesting that parenteral transmission played a role in the spread of HIV in the early epidemic. Since then Libya in cooperation with the European Union has released the Bulgarian nurses and scaled up high efficient scientific research programs to treat the infected victims, introduce a harm-reduction approach and trace the infected individuals (13, 14). The program was found to be successful and has been shown to reduce risky practices and HIV transmission among the Libyan population. A community-based study in the country points out that the number of newly registered HIV cases dropped immensely in Libya, HIV prevalence in the general population stands at >0.05%. Indicating that the epidemiological situation had stabilized (15). This epidemiological stability was disrupted by internal protest in 2011 in the Eastern region complicated by NATO military intervention and a continuous civil war until October 2020 (16,17).

Human migration and population displacement are likely to have an impact on epidemic dynamics within and among the Libyan regions. HIV Infected people move to new places, they might disseminate new viral strains. In addition, new groups of susceptible individuals may be generated, affected by war, with potentially poor access to health care. Hence then an increased export of HIV from the Eastern region (most heavily HIV-affected region and where the war escalation started) to other Libyan regions are expected (18,19). The dual burdens of HIV/AIDS and armed conflict will be a major obstacle to development in the country. However, little is known about the dynamics of the relationship between the two crises in Libya. Hence then, a clearer understanding of the dynamics of the interface between conflict and HIV is crucial for the development of effective and efficient strategies to reduce population risk. The objectives of this study were to analyze the changes in prevalence and dynamics of HIV-1 subtypes overtime during the Libyan conflict and to determine how the viral infections have been redistributed among the Libyan geographical regions?.

## MATERIALS AND METHODS

### Data Collection

In Libya, the HIV surveillance system is based on mandatory, anonymous notification of newly diagnosed HIV cases by laboratories all over the country combined with epidemiological information on the mode of transmission and other clinical data, as reported by physicians and trained clinical epidemiologists. We used the data collected from all over Libyan regions of HIV-infected persons during the Libyan conflict from 2011 till, 2020 as shown in Table 1.

**Table 1.** Number of reportedly HIV-infected people during the Libyan armed conflict in ten years (2011-2020).

| Year | No. Cases* | Male | Female | Ratio |
| --- | --- | --- | --- | --- |
| 2011 | 371 | 293 | 78 | 4:1 |
| 2012 | 486 | 385 | 101 | 4:1 |
| 2013 | 459 | 338 | 121 | 3:1 |
| 2014 | 439 | 359 | 80 | 4:1 |
| 2015 | 511 | 372 | 139 | 3:1 |
| 2016 | 309 | 212 | 97 | 2:1 |
| 2017 | 471 | 382 | 89 | 4:1 |
| 2018 | 492 | 387 | 105 | 3:1 |
| 2019 | 489 | 382 | 107 | 4:1 |
| 2020 | 512 | 399 | 113 | 4:1 |
| Total | 4539 | 3509 | 1030 | 3:1 |

Number of seroincident individuals\*.

### Demographic Factors

Demographic data included were collected for all patients including Gender (Male/Female), Behavioral data and Transmission risk factors, (people who inject drugs (PWID), heterosexuals, others, residential area (region, province postcode), Resident, or displaced, Age category (< 20,20-29, 30-39, 40-49, ≥50).

### Tracing and Migration Pathway

A comprehensive and detailed follow-up of each patient infected with HIV was carried out during the ten years of the Libyan conflict from 2011 to 2020. This consists of patient location, HIV conditions, and the transmission dynamics. The viral gene flow was indicated as the number of migration events; this is the number of viral strain movements from one location (i.e region) to another.

The numbers of HIV-1 subtype migration exportation and importation events were correlated with the epidemiological characteristics of HIV within each region. This includes all the Libyan HIV data set infected patients. Epidemiologic data were gained by reconstruction of the migration route For each region (West, Central, South &East), we looked at the following:

- The numbers of exportation viral strain migration events from one region to all of the other regions.
- The numbers of importation viral strain migration events from one region to all of the other regions.

### Geographic and Statistical Analysis

Viral geographic transition flow that might be associated with the military conflict in the East, where the conflict started was traced all over the country and all cases were officially according to the national case report as previously described(5). Chi-square tests of independence for the association of demographic data with HIV subtype were performed in R v3.6.2 (24) using the gplots and corrplots packages (25,26) and linear regression and correlation coincident were calculated in MATLAB® v2020a and regarded a significant with p values < 0.05 (27)

### Ethical Approval

Routinely collected sequence and demographic data on all newly notified HIV-1 infections are linked and irreversibly de-identified to enable public health research in the country as previously described (15,17). The study was approved by the Libyan National Ethical Committee (Approval No. LY NS, HIV473221). It was conducted under the Helsinki Declaration and the supervision of the Libyan Study Group of Hepatitis & HIV (20, 21).

## RESULTS

A total of 4539 different strains of HIV were available from the Libyan HIV database. These data were collected for all over the Libyan regions (West, East. Middle, and South) within ten years period from the start of the armed conflict 2011 till 2020. Of these reported strains 3509 (77.3%) were reported from males and 1030(22.7%) females (M: F ratio 3:1). The number of reported cases varied from one year to another during the study period. They were increased from 371 in 2011 to reach 512 in 2020 and the reported incidence rate (IR,) (number of reported cases/population) rose from 6.0:100,000 in 2011 to reach up to 9.0:100,000 in 2020 as shown in Table 1.

The demographic, clinical, and HIV-1 sequences data of the enrolled participants were shown in **Table 2**. The median age of participants was 37 years [inter-quartile range (IQR) of 26–49 years]. Of 4539 participants involved in the study, 1369 (30.2%) were from the Eastern region, 1685(37.1%) Western region, 937(20.6%) Middle region, and 548(12.1%) Southern region. A total of 3070 (67.4%) were reported among resident individuals and 1469 (32.4%) from the displaced population. No variation in the number of reported cases within resident individuals. It was found to be 16 % in 2011 and 17.2 % in 2020 while those reported from displaced individuals increased steadily from 2.9% in 2011 to 4.7% (2013-14), 12.6 %, and 7.2 % and then declined to 4.8 % at the end of the conflict period.

**Table 2.**
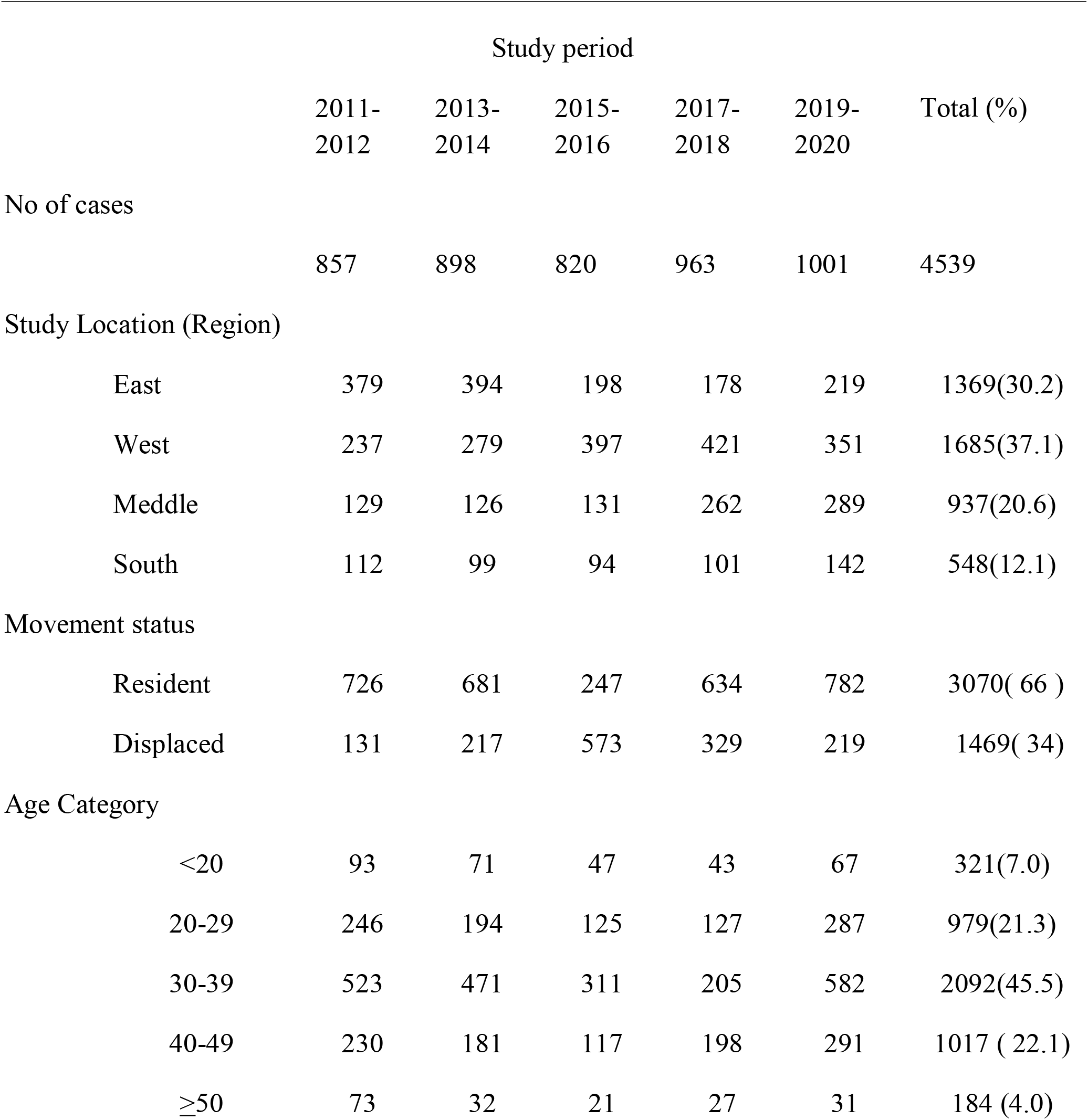
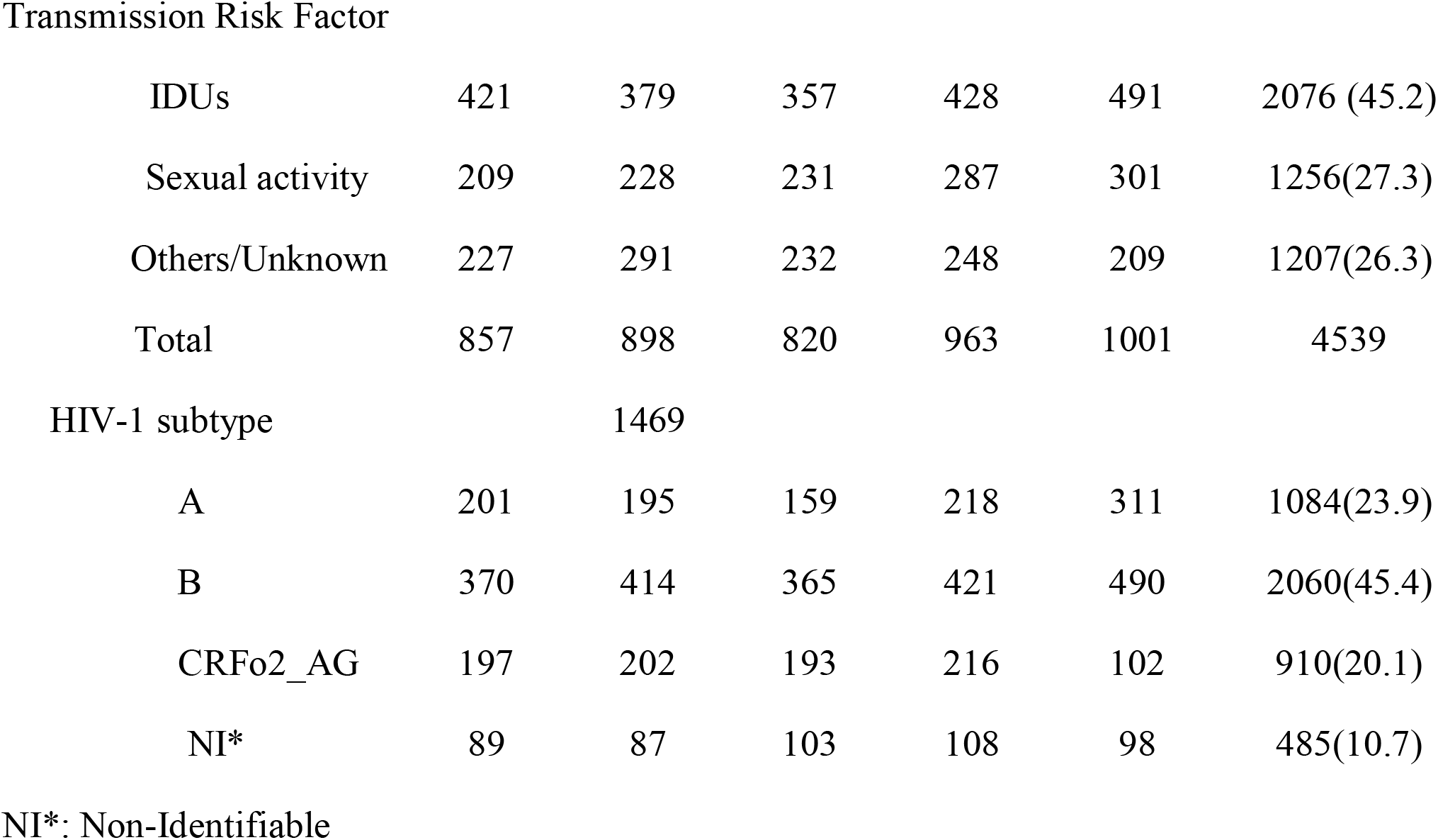
Demographic characteristics of HIV infected population during the Libyan armed conflict-2011-2020

The majority of the study participants attributed to IDUs (Injecting Drug users) accounted for 2076 (44.7%). Followed by those with a high-risk sexual behavior 1256 (27.7%) and 1207 (26.6%) with other risk factors. No significant changes were found for transmission risk factors during the investigation period 2011-2020 (*P*> 0.01). The proportion of IDUs cases was reported to be (9.3%) in 2011 and (10.8%) in 2020 and (5% to 6.6 %) for sexual contacts and 6.4% and 6.4 for other risk factors.

The overall HIV subtype distribution was A (1084(23.9%), B 2060(45.4%), **CRF02_AG 910** (20.1%), and others 485(10.7%). The trends of HIV subtypes were changed overtime during the conflict period. There is a substantial difference in the emergence of each subtype during the ten years. We observed an increase in the proportion of subtype B infections from 8.2% to 10.8% and subtype A from 4.4 % to 6.7%. For **CRF02_AG** we found a significant decrease over time as it varied from 4.3% to 2.2% during 2011-2020.

The transmission dynamics of HIV Type 1 in the Libyan Population during the armed conflict was illustrated in **Figure 1**. The country is classified into twenty-two provinces within the four national regions, the West region (7 provinces), Central region (3 provinces), South region (5 provinces), and East region (7 provinces). According to the geographic data set analysis, the East region was the main exporter accounting on average for 93.5% of the migration events in Libya. The geographic strains movements were mostly observed in this region. The most strongly supported viral migration route was found between Benghazi and Tripoli, Benghazi to Misrata. Other routes connecting Benghazi to Sebha and Western Mountains were frequently reported. Other routes of Genetic flow connected Tripoli to the Western mountain, Misrata to Tripoli, and Misrata to Sebha was also reported.

**Figure 1.**
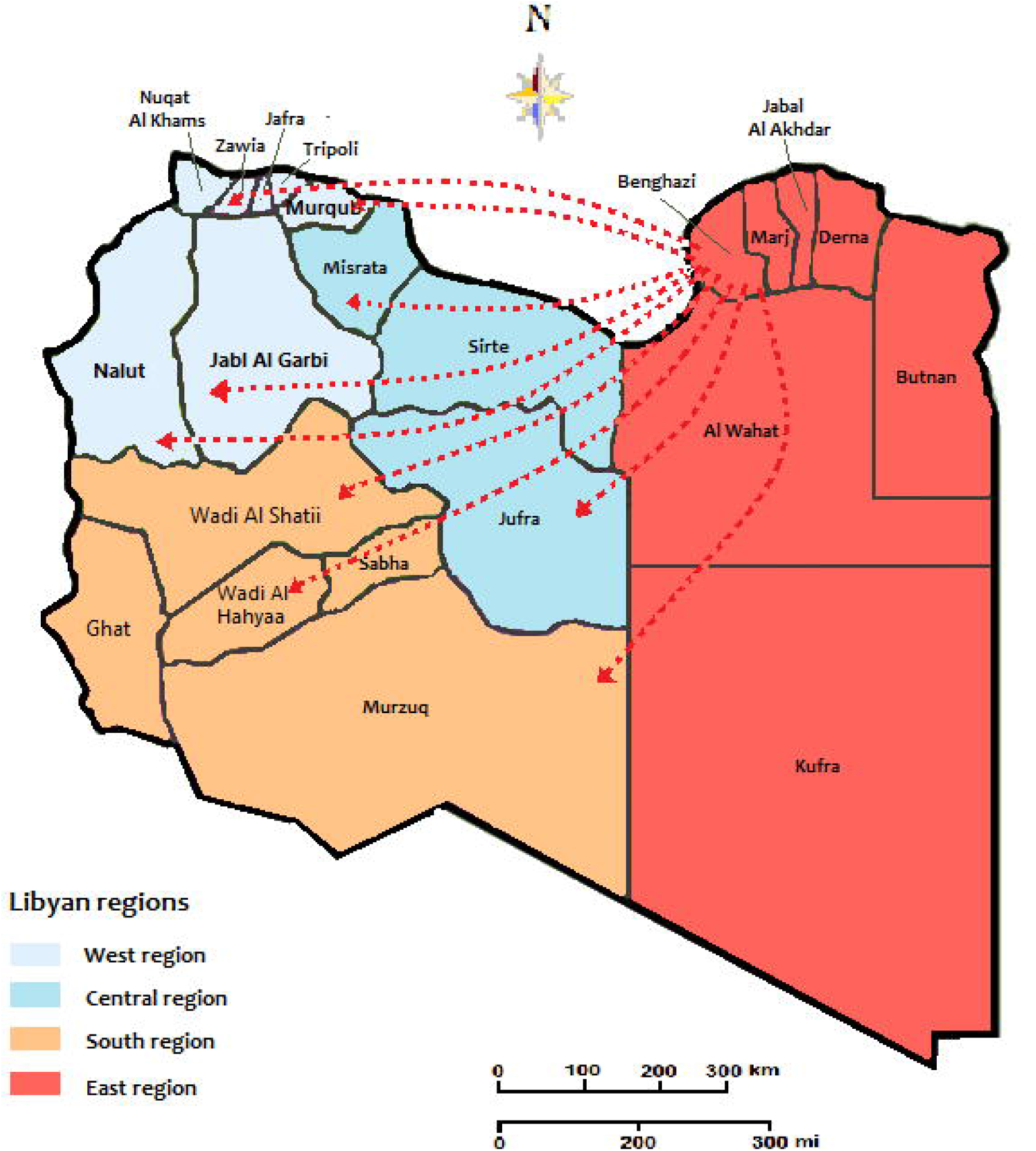
The migration and expansion patterns of HIV cases during the conflict time 2011-2020. The arrows indicate the directionality of virus flow movement from the residential location.

**Table 3** illustrates the different HIV-1 strains that migrated from and to different regions during the armed conflict. A total of 972 type-able strains 504(51.9%) were migrated all over the country. Of these 471 (48.5%) migrated from the Eastern Region including 321(33.0%) recombinant CROF, followed by B 143 (14.7%) and only 7(0.7%)and HIV-1 A. The meddle region received 17 (1.7%) migrated stains (9 CROF, 6A 2B), followed by Western Region 9 (0.9%) strains and only 7 (0.7%) Southern Region.

**Table 3.** Inter-location series of HIV-1 strains in Libya during the armed conflict 2011-2010

| No cases(%) | Migrated to (%) |  |  |  | Migrated from (%) |  |  |  |
| --- | --- | --- | --- | --- | --- | --- | --- | --- |
| Location | Total | A | B | CROF | Total | A | B | CROF |
| West Region |  |  |  |  |  |  |  |  |
| Tripoli | 319 | 13 | 224 | 82 | 9 | 1 | 5 | 3 |
| Meddle Region |  |  |  |  |  |  |  |  |
| Misrata | 107 | 09 | 75 | 23 | 17 | 2 | 06 | 09 |
| East Region |  |  |  |  |  |  |  |  |
| Benghazi | 39 | 07 | 19 | 13 | 471 | 7 | 143 | 321 |
| South Region |  |  |  |  |  |  |  |  |
| Sebha | 03 | 0 | 02 | 01 | 07 | 4 | 3 | 0 |

Furthermore, 468 (48.1%) strains were migrated to different regions including, 319 (32.8 %) migrated to The western region particularly Tripoli, including B 224(23%) C 82 (8.4%) A13 (1.311%), followed by Meddle region 107 (%) A 09(9.0%) B 75(7.7%) C 23(2.4%).Although only 39 (4.0%) and only 3 Strains migrated to Eastern and southern regions respectively.

The prevalence of HIV and the distribution of multiple HIV subtypes in different geographic locations during the Libyan conflict were presented in **Figure 2**. In the first five years period (2011-2015 -Figure 2A), the Eastern region showed the highest prevalence of HIV particularly in Benghazi estimated to be> 0.8% followed by Marj, Darna, and Butnan and to less extent Al Wahat and Kufra. In these provinces, CROF was the predominant circulating strain accounted for 60% followed by Strain B with 20%. In the West region, Tripoli reported a HIV prevalence of 6% followed by Nalut 4% others are less than 0.2%. In the Western provinces, HIV-B was the predominant strain which has reached 50% followed by HIV-A and CROF which accounted for 20% each. In Central province, HIV prevalence was reported to be 0.6% in Misrata followed by Sirte and Jufra. HIV –B and CROF were the predominant circulating strains, where they occupied 45 % and 25%respectivelly. In the South region the HIV prevalence was reported to be very low (<0.4%). The predominant circulating strains in these provinces were B(40%,), A(30%), and to less extent CROF(20%).

**Figure 2.**
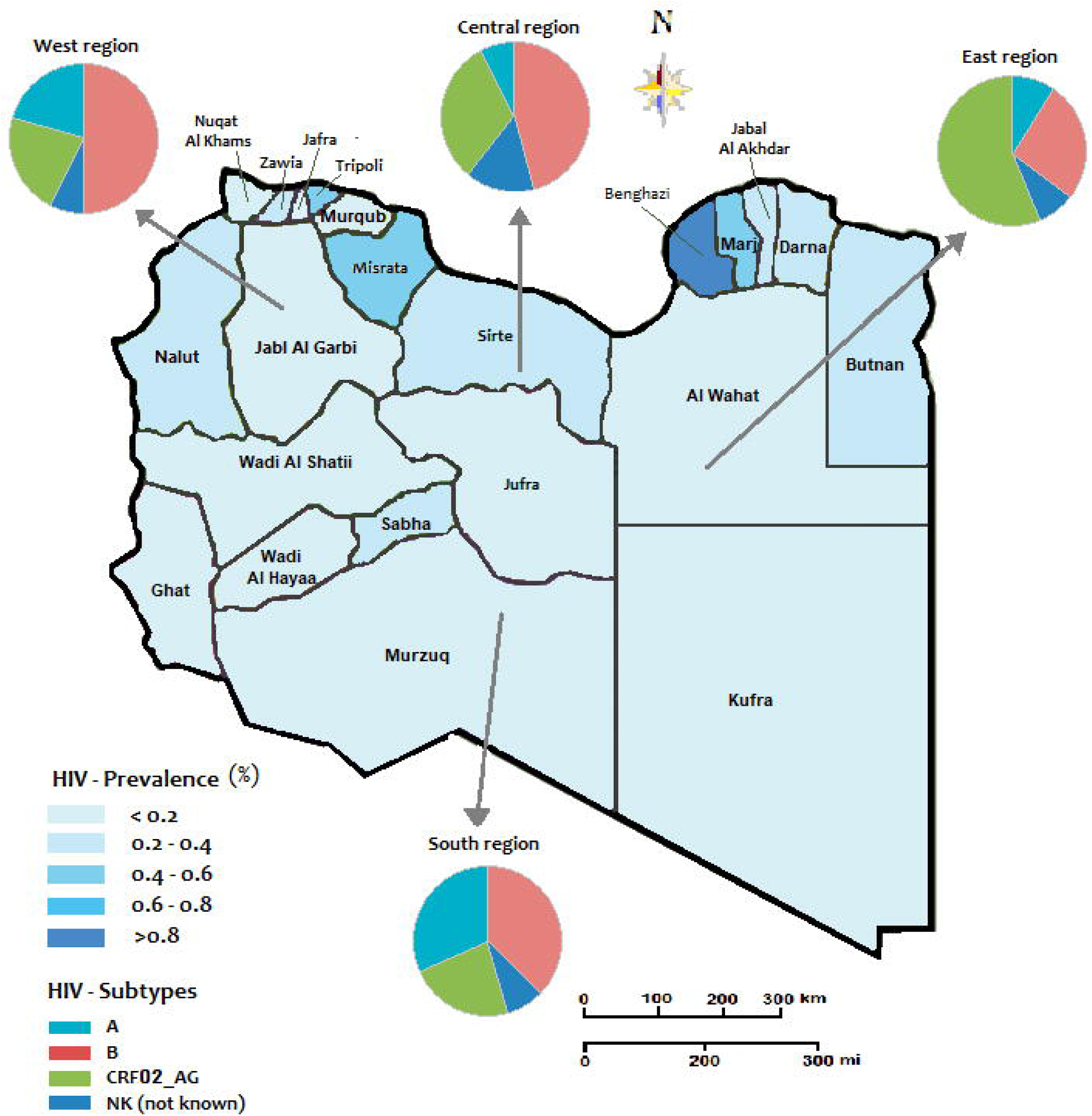
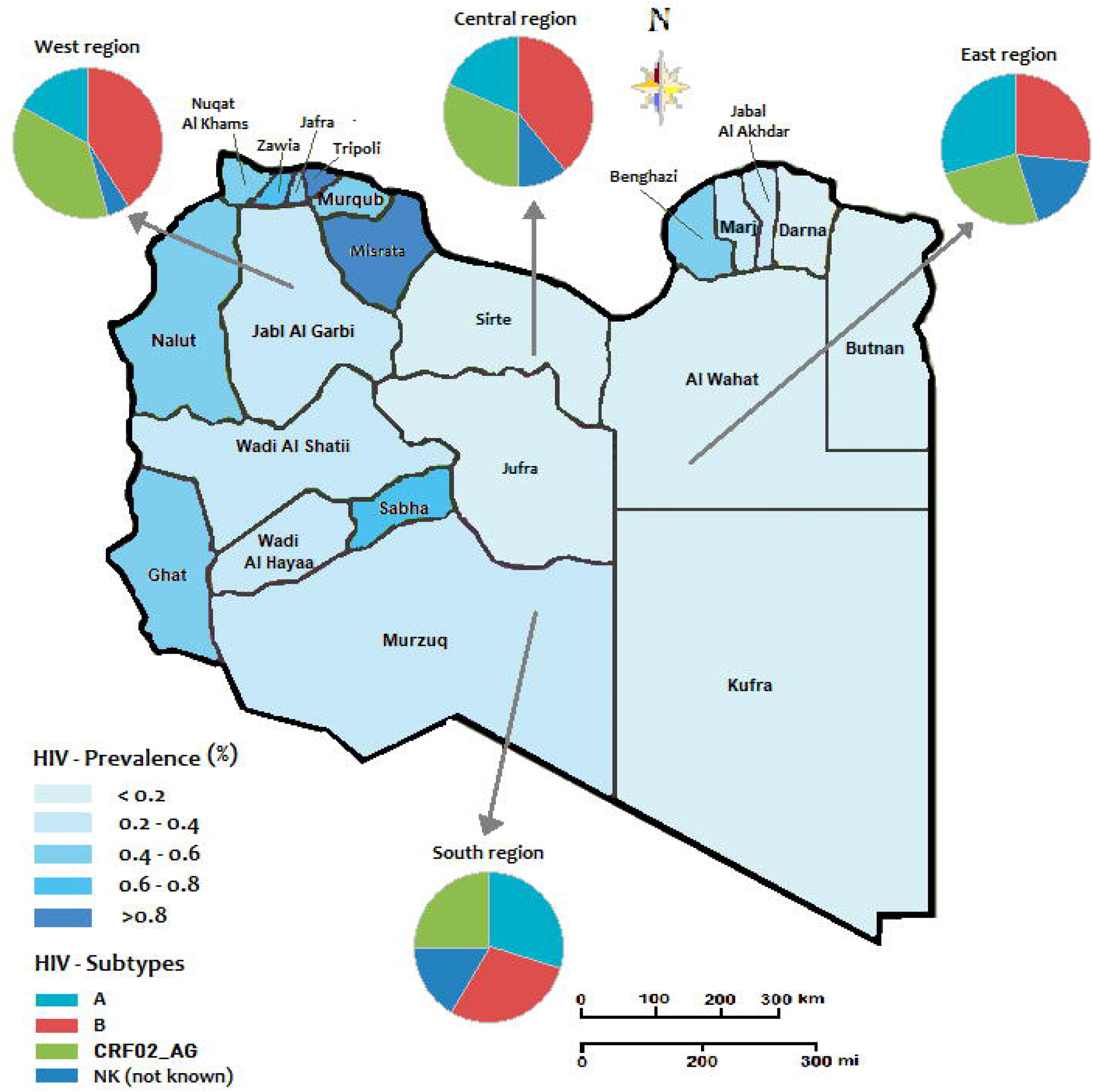
The regional prevalence of HIV and geographic distribution of HIV-1 subtypes during the Libyan conflict A- the period from 2011 to 2015 B- period 2016 to 2020

**Figure 3.**
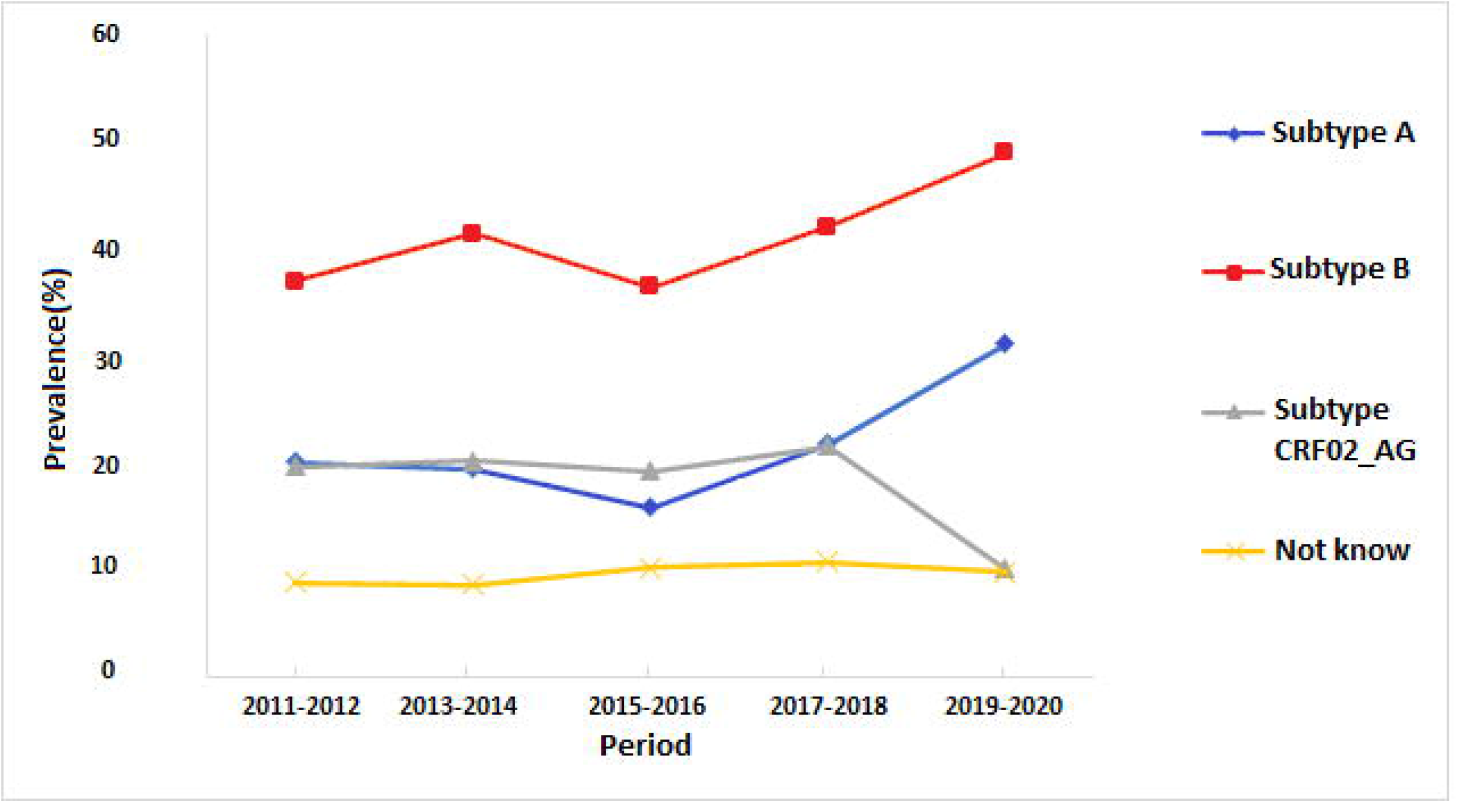
Trends of the prevalence of different HIV-1 subtypes in Libya during the armed conflict-2011-2020

In the second period of the conflict (2016-2020) Figure 2B. The Western region showed a higher HIV prevalence. The highest prevalence was reported in Tripoli, Zawia, Nuqat Al-Khams, Jafra,Murqub, Nalut, and to less extent Jabl-Al Garbi. In these provinces, the highest-circulating HIV strains were B (40%) CROF (35%), and A (20%). In the Central region the highest prevalence was reported in Misrata (>0.8%) and only < 0.2% in Sirte and Jufra. HIV-strain B accounted for 40% of the circulating strains followed by CROF (35%) and B (20%). In the Southern provinces, the HIV prevalence reached 0.6% in Sabha and Ghat and 4% in the other three provinces. The most circulating strains occupy the Southern provinces were Strains A&B accounted for 30% each followed by CROF (25%). In the Eastern region HIV prevalence was 0.6% in Benghazi and < 0.2 in the other provinces. Furthermore, the most predominant circulating strains were B (25%), CROF (25%), and A (20%).

## DISCUSSION

Armed-Conflicts may have profound effects on the spread and dissemination of the HIV pandemic which should be considered. A long period of conflicts has resulted in greater naïveté regarding the epidemiology and prevention of the disease in Sub-Saharan and North African countries (22). In this study, we investigated the dissemination and spread of HIV during the armed conflict in Libya by analyzing the spatial and temporal patterns of the viral flow during the ten years of the conflict. Our data indicated that HIV-reported strains have moved from the regions involved in the armed -conflict to the rest of the country. The Eastern region was the most exporter of HIV strains, as it appears that the strains were originated directly or indirectly from Benghazi; the largest city in the East region. The east location is heavily affected by HIV before and at an early stage of the armed conflict. Within the period of 2011-2014, a total of 1755 cases were reported in the eastern region accounting for 38.7% of all nationally registered cases of HIV infection.

Demographic analysis in this study indicated that the highest HIV prevalence was reported among IDUs accounting for 2076 (45.2%) cases followed by sexual activities 1256 (27.7%) cases. This was mainly common among the middle age group of (30-39) years followed by 40-49). Though it was less among other age groups. The genetic analysis has shown HIV-1 subtype A accounted for 2060(45.4 %) cases followed by HIV-1 subtype A 1084(23.9%) and to less extent CRF02_AG, for 910 (20.1%). As IDUs are a main contributing factor among war internal displacement in the study. This may create a favorable environment for new HIV transmission and facilitate the generalization of the HIV epidemic in Libya. Similar studies on HIV in conflict-affected settings in Africa suggest that regions that have received new imported HIV strains also have appropriate conditions for those strains to flourish (23,24).

The prevalence of HIV and the spread of HIV-1 subtypes have experienced tremendous changes during the Libyan armed conflict. Our study has shown the inter-location of HIV strains within the Libyan regions is an emerging phenomenon. The number of viral exportation correlates well with the number HIV infected patients in each region. We have observed more infected people moved from the locations from which the virus is originating. This is evident in our study as more HIV-infected individuals moved from the main exporting locations (this is Benghazi at Eastern region) to the main importing locations particularly at the Western region (i.e Tripoli) and Meddle region (Musrta). This inter-location has resulted in a major change in the prevalence of HIV infection and the dissemination of HIV strains within the Libyan region. Before and during the early stage of the conflict the HIV prevalence was similar among most of the Libyan regions. While it has been geographically inter-located since then. In the Eastern region where the conflict has exsanguinated, it was changed drastically from 0.6-0.8 % in (2011-2015) to 0.2-0.4 % (2016-2020). Although the western and Meddle region was escalated from 0.2-0.6% (2011-2015) to 0.4 - 0 >0.8 during 2016-2020. With no significant change in the national prevalence of HIV and HIV strains during the conflict period (2011-2020).

Population displacement is a major consequence of armed conflict which reflects greatly on the health of displaced people. In Libya, over 250,000 people were displaced in 2011 this was further increased as the armed conflict escalated from 2014 to 2020. This could be reflected in the epidemiological patterns of microbial disease and health conditions of the displaced people including HIV Infected individuals (25,26).). In this study, major transition patterns of the viral strains movements during the armed conflict have been reported. HIV-subtype CRF02_AG; the main strain resident in the East region and rarely reported in the other Libya regions before 2011 accounted for 68% of the migrated strains to the other regions particularly the west and central regions. The number of the exported strains correlated well with the number of registered HIV cases in each region. More infected people have moved away from the conflict zone to a more secured area. This reflects the redistribution of preexisting infections within the country. Many resident people who were initially infected in the eastern region have moved to central and meddle regions due to war, resulting in a redistribution of HIV viral strains within the country. This is in concordance with other data reported from other countries inflated by war such as Uganda and Ukraine(27,28). Furthermore, population displacement may be reflected in the treatment of HIV-infected individuals as patients who had to relocate because of the conflict may be more likely to reduce treatment adherence. Hence then the spread of HIV-1 strains is becoming a serious public health challenge that requires further studies to understand deeply their origin and distribution (29).

Different comments should be raised concerning the limitations of this study. First, the study is carried during the conflict time where not all the needed data are easily available or accurately reported. Second, our data is based on the officially reported cases of HIV-infected patients and the number of typed stains. Hence then caution is advised regarding the exact number of HVI infected cases in the present study which may not be representative of the real HIV population in the country. Hence then further studies are needed at the post-conflict period to improve the accuracy of the number of HIV migrant strains that fit the population distribution with the Libyan regions (30,31). However, this study indicated clearly that human migration dynamics may play a key role in the dissemination of new viral strains and influenced the risk of HIV. This may exacerbate vulnerability and accelerates the spread of HIV in post-conflict settings. Such juxtaposition which is evident in this study has already been reported to some extent in certain African countries including Mozambique(32). Unfortunately, the international community that intervened in 2011 and initiated this armed conflict, has compartmentalized its responses to HIV and conflict in Libya. Therefore, efforts should be combined and studies are needed to formulate long-run policies to precludes an integrated and aggressive attack on the HIIV in Libya (33,34).

## CONCLUSION

This is the first study to investigate the impacts of the armed conflict on the epidemiological patterns of HIV infection in Libya using spatial and temporal analysis. Our findings support the initial spread of HIV infections and HIV strains providing quantitative measurements of the spread and dissemination of the infection at regional and national levels. These findings contribute greatly to the fundamental understanding of the patterns and transmission dynamics of HIV during the armed conflict. The migration of HIV strains represents an enormous surveillance challenge. Hence then national intervention policies during and at post-conflict periods should be implemented considering such understanding (35,36).

## Data Availability

Here in we confirm that the data available free

## DATA AVAILABILITY STATEMENT

The data presented in this paper are freely available upon request

## AUTHOR CONTRIBUTIONS

MD conceived and designed the study, wrote the paper, designed the analysis, analyzed the data, and performed cartography. MD and AE-B contributed to the analysis tools. AE-B and MA made substantial contributions to conception and design, acquisition of data, or analysis and interpretation of data. MD, MA, and AE-B provided advice and critically reviewed the manuscript. All authors read and approved the final manuscript

## ACKNOWLEDGMENTS

We are deeply grateful to the Libyan Study Group of Hepatitis & HIV and the Department of Medical Microbiology, Faculty of Medicine, University of Tripoli, for their help and support.

